# Association between ABO blood groups and clinical outcome of coronavirus disease 2019: Evidence from two cohorts

**DOI:** 10.1101/2020.04.15.20063107

**Authors:** Xianfei Zeng, Hongyan Fan, Dongxue Lu, Fang Huang, Xi Meng, Zhuo Li, Mei Tang, Jing Zhang, Nannan Liu, Zhixin Liu, Jingya Zhao, Wen Yin, Qunxing An, Xijing Zhang, Xingbin Hu

**Affiliations:** Department of Laboratory Medicine, Shaanxi Corps Hospital, Chinese People’s Armed Police Forces, Xi’an, China; Department of Laboratory Medicine, Huoshenshan Hospital, Wuhan, China; Department of Blood Transfusion, 940 Hospital, Lanzhou, China; Department of Anesthesiology and Perioperative Medicine, Xijing Hospital, Xi’an China; Department of Laboratory Medicine, Xi’an Chest Hospital, Xi’an, China; Department of Laboratory Medicine, First Affiliated Hospital of Xi’an Medical University, Xi’an, China; Intensive Care Center, Xijing Hospital, Xi’an, China; Intensive Care Unit, Huoshenshan Hospital, Wuhan, China; Department of Blood Transfusion, Xijing Hospital, Xi’an, China; Department of Blood Transfusion, Tangdu Hospital, Xi’an, China

**Keywords:** COVID-19, ABO blood type, susceptibility, ARDS, AKI, mortality

## Abstract

Severe acute respiratory syndrome coronavirus 2 (SARS-CoV-2) has become the third most common coronavirus that causes large-scale infections worldwide. The correlations between pathogen susceptibility and blood type distribution have attracted attention decades ago. The current retrospective study aimed to examine the correlation between blood type distribution and SARS-CoV-2 infection, progression, and prognosis in patients with coronavirus disease 2019 (COVID-19). With 265 patients from multiple medical centers and two established cohorts, we found that the blood type A population was more sensitive to SARS-CoV-2. Moreover, the blood type distribution was not relevant to acute respiratory distress syndrome (ARDS), acute kidney injury (AKI), and mortality in COVID-19 patients. These findings are indicative of coping with the great threat since it probed the relationship between blood types and ARDS, AKI, and mortality, in addition to susceptibility in COVID-19 patients.

## Introduction

In the late part of 2019, a cluster of unexplained pneumonia cases occurred. Patients with fever, cough, and fatigue were diagnosed with a novel coronavirus disease, officially named as COVID-19 by the World Health Organization in February 2020. Severe acute respiratory syndrome coronavirus 2 (SARS-CoV-2) has become the third coronavirus that causes large-scale infections among humans, following severe acute respiratory syndrome coronavirus (SARS-CoV) and Middle East respiratory syndrome coronavirus in the past two decades. The coronavirus outbreak has caused 81,008 infections in China by March 20, 2020 and spread to 171 other countries. More than 1,000,000 cases of COVID-19 have been reported globally, among which more than 50,000 people died. The severe outbreak of COVID-19 made it a public health threat all over the world. All nations are allocating numerous medical and scientific resources to fight against the COVID-19 pandemic.

Scientists have been investigating the pathology, epidemiology, clinical characteristics, diagnosis, and treatment of COVID-19. It is meaningful to perform relevant research on the susceptibility and outcome of COVID-19. Chen et al. proposed that older men with comorbidities might develop severe symptoms and have poor prognosis by conducting a retrospective, single-center study of 99 patients [1]. The data indicated that gender, age, and physical condition might be risk factors of COVID-19 outcomes. With regard to SARS in 2003, Cheng and colleagues showed an association between ABO blood group and SARS-CoV infection after exposure [2]. The blood group O population was believed to be less susceptible to SARS. The inherent mechanisms were also investigated, suggesting that human anti-histo-blood group antibodies could block the interaction between the viruses and cells by binding to the spike (S) protein [3]. These results indicate that various factors can affect the spread of coronavirus disease, including blood groups.

Antigens in the blood group system are expressed not only on the surface of human red blood cells, but also on many tissues [4], which extends the clinical significance of these antigens beyond compatible blood transfusion. Studies on the relationship between the blood group system and human health started in the early 1900s. Blood groups have been associated with the occurrence of several diseases, including cardiovascular disorders, neurological disorders, cancer, and infections [5-6]. Previous studies have demonstrated that the non-O blood group was associated with increased risk of coronary heart disease through a meta-analysis, which might be due to the high level of von Willebrand factor (vWF) in plasma [7-9]. Recently, a study of 406,755 unrelated individuals was published by a research group from the Netherlands [10]. The data indicated that ABO blood group was associated with 11 health problems, especially cardiovascular outcomes. Meo et al. analyzed 15 publications from a public database and found that blood group B was associated with a high risk of type 2 diabetes, while Rh blood groups seemed to have no effect [11]. In addition, Duffy blood group antigens have been shown to be receptors for plasmodium vivax invasion into the red blood cells, which explains the high ratio of the Fy(a-b-) genotype of people in Africa [12]. Overall, these studies suggested that the blood group system was associated with the occurrence of several diseases.

The relationship between blood groups and human diseases is more than a causal association of incidence. Blood groups may influence the progress and outcomes of diseases. The blood type O affects the volumes of grey matter in the cerebellum, which might be associated with the decline in cognitive function [13]. A retrospective study was conducted by a Japanese group to analyze the impact of ABO blood group on outcomes in patients with severe trauma. The results suggested that blood type O was associated with high mortality and less ventilator-free days [14]. Moreover, blood groups have been shown to be relevant to the development and outcomes of chronic heart failure [15] and esophageal squamous cell carcinoma [16]. However, other studies provided conflicting evidence that blood groups were not risk factors for several diseases [17-18].

To examine the correlation between blood type distribution and SARS-CoV-2 infection, progression, and prognosis of COVID-19, the current study conducted a retrospective investigation of patients from multiple medical centers.

## Materials and methods

### Study design

#### Study populations

Two retrospective cohorts diagnosed with COVID-19 were enrolled in this study between February 5 and March 20, 2020. The first cohort comprised patients with mild symptoms admitted in three hospitals in Xi’an, Beijing, and Wuhan. The second cohort comprised critically ill patients from the two hospitals in Xi’an and Wuhan. Five hospitals in this study were assigned by the government as the treatment centers for COVID-19 patients. All study participants were of Chinese Han nationality population and aged >13 years. In the critical cohort, patients were excluded if they died within 24 h upon admission or had current evidence of congestive heart failure, myocardial infarction, or severe isolated head trauma, which was not related to the progress of COVID-19. The study was approved by the Institution Review Board of the Shaanxi Province Corps Hospital of Chinese People’s Armed Police Forces, and oral informed consent was obtained from all patients or first-grade relatives.

#### Diagnosis and outcome definitions

COVID-19 was defined according to the World Health Organization interim guidance [19]. COVID-19 was confirmed by a positive real-time reverse transcriptase polymerase-chain-reaction test of SARS-CoV-2 on nasal and pharyngeal swab specimens from suspected patients. Critically ill patients were defined as those admitted to the intensive care unit (ICU) who required mechanical ventilation or had a fraction of inspired oxygen (FiO_2_) of ≥60% [20-21]. Identification of critically ill patients was achieved by reviewing and analyzing the admission logs and histories from all available electronic medical records. AKI was diagnosed if the serum creatinine levels change within 2 days based on the Acute Kidney Injury Network (AKIN) criteria [22]. Patients were diagnosed with ARDS based on the 2012 Berlin definition [23].

#### Laboratory testing and data collection

ABO blood type of patients was determined by standard RBC typing performed for clinical purposes. Nucleic acid testing was performed according to the Technical Guidelines for SARS-CoV-2 Laboratory Testing (from the third to fifth edition), Chinese version, issued by the National Health Commission of the People’s Republic of China.

Demographic data such as age, sex, and medical records including results of laboratory testing, chest radiographs, medical history, and clinical outcomes were collected from all participants. The clinical outcomes (e.g., development of ARDS or AKI, discharged, and mortality) were monitored until the last date of follow-up, March 20, 2020.

### Statistical analysis

Continuous variables were expressed as median (25th and 75th percentiles), while categorical variables were expressed as percentages. The Mann–Whitney U-test or the Kruskal–Wallis test, as appropriate, were used to compare the medians of continuous variables between or among groups. Categorical data were tested using a χ^2^ test or Fisher’s exact test. The unadjusted associations of ABO blood type and SARS-CoV-2 infection, development of ARDS or AKI, and death were tested separately among the cohorts using Pearson’s χ^2^ test. Odds ratios (ORs) and 95% confidence intervals (CIs) were calculated to describe the possible effect on the occurrence and development of COVID-19.

Given the limitations of a single factor analysis, we used multivariable logistic regression to further adjust the association of ABO blood type with occurrence of ARDS, AKI, or death for potential confounders, including the common risk factors of age, sex, pre-existing pulmonary disease, history of cardiovascular disease, diabetes mellitus, hypertension, and chronic kidney disease in the critical cohort. The history of diabetes mellitus was selected as a crucial confounder that needs to be adjusted because diabetes was reported to be protective against the development of ARDS, but as a risk factor for AKI and death. Most importantly, it has been associated with the ABO blood group [24]. All baseline variables listed in Table 4 that had an unadjusted association with ARDS, AKI, death, or ABO blood type at p<0.20 were considered for inclusion in the multivariable models. Additional potential confounders were only included in the final multivariable model if they altered the unadjusted OR for the association between ABO blood type and clinical outcomes by >10% in the bivariate analysis [25].

For statistical presentation, each blood group was compared with a reference group (blood type O or B). A stratified analysis of a reported region was not performed given that there were no substantial differences in allele distributions and divergent genetic background in participants of Han nationality between different regions. Statistical analyses were performed using SPSS (version 22.0) for Windows. A p-value of <0.05 was considered significant.

## Results

### Characteristics and outcomes of the study cohorts

In the current study, we enrolled 137 patients with mild pneumonia and 97 patients with severe pneumonia in the mild and critical cohorts, respectively, (Figure 1). The demographic data, clinical characteristics, and incidence of ARDS, AKI, and death of the enrolled patients with different blood types are presented in Tables 1 and 2. In this study, the individuals with AB blood type were not statistically analyzed because of the insufficient number of patients (14 in the mild cohort and 5 in the critical cohort). In addition, individuals with blood type AB only accounted for 9.0% of the Chinese population [26].

**Table 1.** Demographic data and clinical characteristics of mild cohort patients.

| **Characteristics** | **A type**<br>**(n=54)** | **B type**<br>**(n=34)** | **O type**<br>**(n=49)** | ***p* value** | **All patients**<br>**(n=137)** |
| --- | --- | --- | --- | --- | --- |
| Age (years) | 56 (49, 65) | 50 (40, 62) | 48 (36, 57) | 0.187 | 52 (40, 64) |
| Age ≥ 60 years | 21 (38.9%) | 10 (29.4%) | 19 (38.8%) | 0.613 | 50 (36.5%) |
| Male | 31 (57.4%) | 18 (52.9%) | 25 (51.0%) | 0.726 | 74 (54.0%) |
| Diabetes | 5 (9.3%) | 4 (11.8%) | 4(8.2%) | 0.857 | 13 (9.5%) |
| Hypertension | 6 (11.1%) | 8 (23.5%) | 8 (16.3%) | 0.303 | 22 (16.1%) |
| Stay in isolation wards (days) | 27 (17, 35) | 26 (12, 38) | 22 (15.32) | 0.629 | 25 (14, 33) |

Data are shown as n (%) for categorical variables and median (interquartile range) for continuous variables.

**Table 2.** Demographic data and clinical characteristics of critical cohort patients.

| **Characteristics** | **A type**<br>**(n=40)** | **B type**<br>**(n=30)** | **O type**<br>**(n=27)** | ***p* value** | **All patients**<br>**(n=97)** |
| --- | --- | --- | --- | --- | --- |
| Age (years) | 68 (57, 74) | 65 (58, 73) | 67 (61, 75) | 0.608 | 67 (57, 75) |
| Age ≥ 60 years | 28 (70.0%) | 22 (73.3%) | 21 (77.7%) | 0.780 | 71 (73.2%) |
| Male | 23 (57.5%) | 19 (63.3%) | 17 (63.0%) | 0.854 | 59 (60.8%) |
| Diabetes | 10 (25.0%) | 6 (20.0%) | 10 (37.0%) | 0.330 | 26 (26.8%) |
| Hypertension | 17 (42.5%) | 15 (50.0%) | 15 (55.6%) | 0.565 | 47 (48.5%) |
| Chronic kidney disease | 2 (5.0%) | 2 (6.7%) | 4 (14.8%) | 0.334 | 8 (8.2%) |
| Cardiovascular disease | 13 (32.5%) | 12 (40.0%) | 5 (18.5%) | 0.207 | 30 (30.9%) |
| Chronic lung disease | 6 (15.0%) | 1 (3.3%) | 1 (3.7%) | 0.128 | 8 (8.2%) |
| Clinical outcomes |  |  |  |  |  |
| ARDS | 25 (62.5%) | 22 (73.3%) | 18 (66.7%) | 0.634 | 65(67.0%) |
| AKI | 7 (17.5%) | 7 (23.3%) | 6 (22.2%) | 0.813 | 20 (20.6%) |
| Death | 8 (20.0%) | 7 (23.3%) | 6 (22.2%) | 0.942 | 21 (21.6%) |

Data are shown as n (%) for categorical variables and median (interquartile range) for continuous
variables. ARDS, Acute respiratory distress syndrome; AKI, Acute kidney injury.

**Figure 1.**
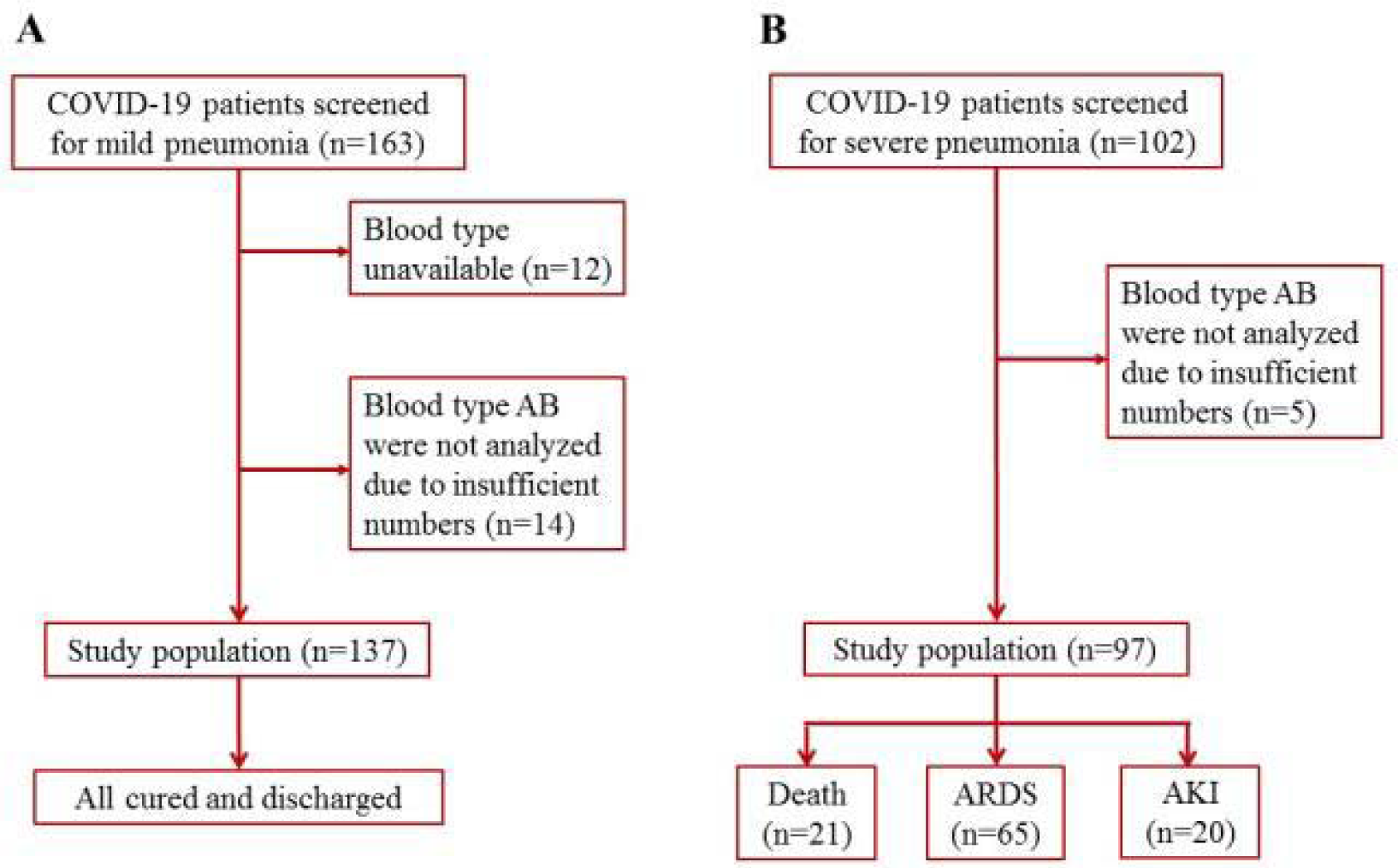
Patients inclusion and exclusion scheme. Patients with mild and severe pneumonia were enrolled in the cohorts, respectively. A. The mild cohort; B.The critical cohort.

In all 234 patients, 133 (56.8%) sufferers were men. When taking age into account, we noticed 67 (57-75) years of age in the critical cohort, and most of them (73.2%) were >60 years of age, but 52 (40-64) and 36.5% in the mild cohort, suggesting that aged sufferers are inclined to develop into the critical ill stage.

All patients in the mild cohort recovered and were discharged. Approximately 67% of patients diagnosed with severe pneumonia developed ARDS. In critically ill patients, 21 (21.6%) died due to multiple organ failure. There was no difference in the constituent ratios of patient characteristics and outcomes among blood types A, B, and O in the two independent cohorts. Overall, the difference between baseline variables was not statistically significant in the two cohorts in terms of blood type distribution.

### Frequencies of ABO blood in COVID-19

We then investigated the potential susceptibility of individuals with different ABO blood types to COVID-19. Table 3 shows the distribution features of the ABO blood types in the mild and critical cohorts. The proportion of blood group A in patients with COVID-19, being 35.76% in the mild cohort and 39.22% in the critical cohort, was significantly higher than that in the reference population (p=0.000 and p=0.005, respectively), which comprised half a million Han Chinese individuals from China [26]. These results imply that the blood group A population may be more vulnerable to SARS-CoV-2 infection.

**Table 3.** Frequency and odds ratio of ABO blood types in mild and critical cohort.

| Blood type | Mild cohort |  |  | Critical cohort |  |  | Chinese population (%) |
| --- | --- | --- | --- | --- | --- | --- | --- |
|  | Frequency (%) | OR(95% CI)* | *p* value | Frequency (%) | OR(95% CI)* | *p* value |  |
| A type | 35.76 (n=54)<sup>#</sup> | 1.40 (1.01, 1.96) | 0.045 | 39.22 (n=40)<sup>§</sup> | 1.63 (1.10, 2.42) | 0.015 | 28.39 |
| B type | 22.52 (n=34) | 0.70 (0.48, 1.03) | 0.066 | 29.41 (n=30) | 1.00 (0.66, 1.54) | 0.986 | 29.33 |
| O type | 32.45 (n=49) | 0.97 (0.69, 1.36) | 0.845 | 26.47 (n=27) | 0.72 (0.47, 1.13) | 0.149 | 33.20 |

OR, odds ratio; CI, confidence interval; \*which compared with the corresponding non-A, non-B, or non-O blood type data referred to Chinese population. The frequencies
for the Chinese population are estimated based on previous literature<sup>[26]</sup>. <sup>#</sup>*p*=0.000, <sup>§</sup>*p*=0.005, compared with the proportion of type A in Chinese population.

**Table 4.**
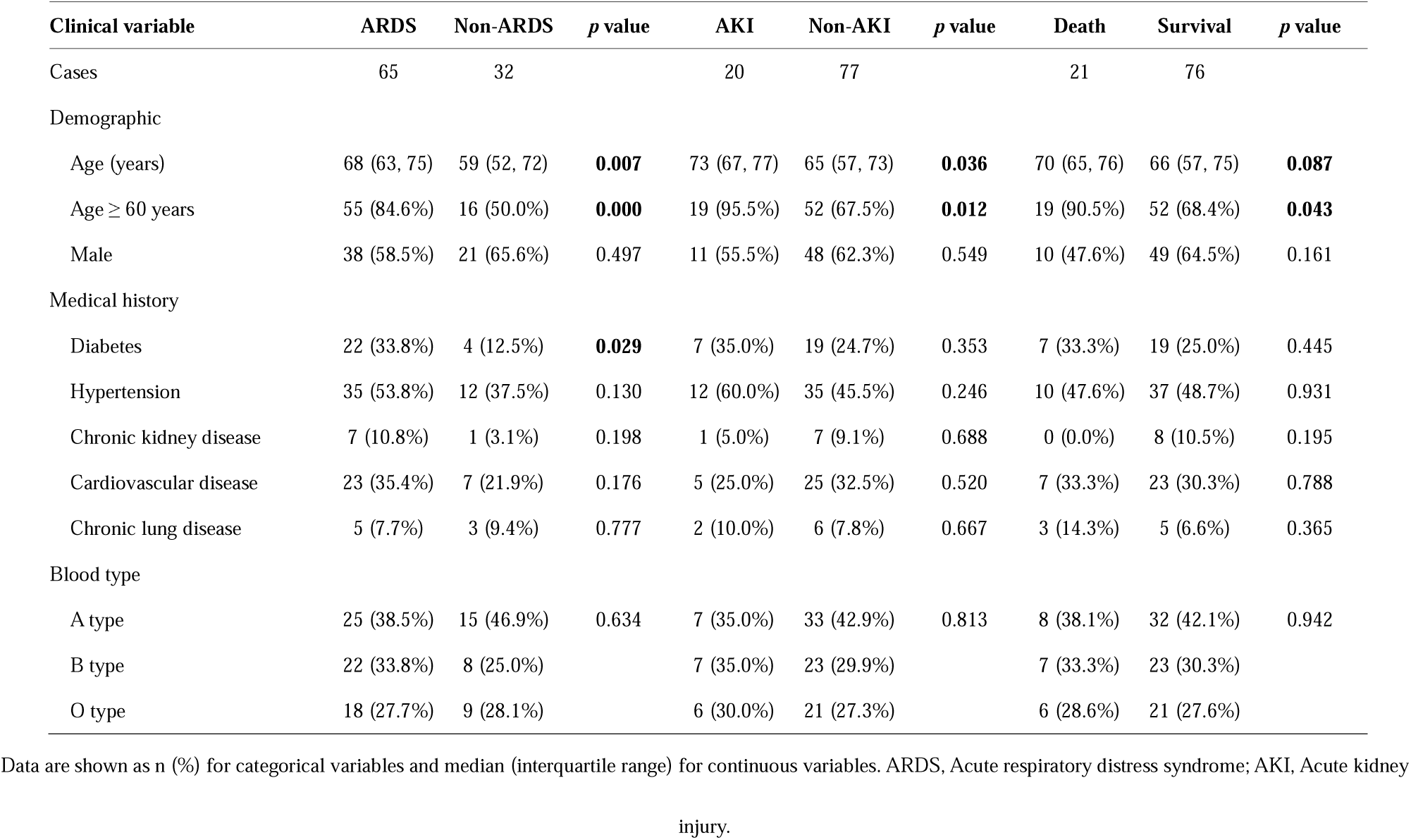
Critical cohort demographics and clinical characteristics based on clinical outcomes.

In addition, blood type A was associated with increased risk of SARS-CoV-2 infection compared with the non-A blood type, with an OR value of 1.40 (1.01-1.96) and 1.63 (1.10-2.42) in the mild and critical cohorts, respectively. However, similar results were not obtained for B and O blood types. These observations further demonstrated that the blood group A population may be more susceptible to SARS-CoV-2 infection.

Compared with the frequency of O blood type in the mild cohort (32.45%) and in the Chinese population (33.20%), the distribution was lower in the critical cohort (26.47%). Although the OR value in the blood type O group relative to non-O blood type was not significantly decreased because of the limited number of enrolled patients (OR=0.72, p=0.149), these findings suggest that the blood type O population did not easily progressed to the critically ill stage.

### Association between blood type and clinical outcome

We further explored the relationship between blood type and clinical outcomes of critical patients with severe pneumonia caused by SARS-CoV-2. Demographics and clinical variables of patients with different clinical outcomes in the critical cohort are shown in Table 4. The age distribution of patients was statistically significant between the ARDS and non-ARDS, AKI and non-AKI, death, and survival groups, thus suggesting that it is closely related to the clinical outcomes and should be included in the logistic model. Although no difference was observed in the gender and the most medical histories between groups, all baseline variables presented in Table 4 were included in the multivariable logistic regression analysis, given their potential influence on patients’ clinical outcomes.

In the multivariable logistic regression analysis, which was used to adjust the association of ABO blood type with clinical outcomes for potential confounders, blood type A was not associated with a higher risk for ARDS, AKI, and death relative to blood type O among patients in the critical cohort (Table 5). Comparisons between blood type A and blood type B did not present any statistically significant associations in the logistic model (data not shown). These data indicate that the blood type of SARS-CoV-2-infected patients is not a potent factor that influences the development and outcome of COVID-19. When the age of patients (<60 and ≥60 years of age) was converted into an ordinal category, results showed that there was an association with ARDS (OR: 6.22 [95% CI: 2.32-16.71]) or death (OR: 9.23 [95%CI: 1.17-72.85]) when adjusted for age.

**Table 5.** Association of blood type with ARDS, AKI, and death risk in critical patients with COVID-19.

| **Blood**<br>**type** | **ARDS** |  |  |  | **AKI** |  |  |  | **Death** |  |  |  |
| --- | --- | --- | --- | --- | --- | --- | --- | --- | --- | --- | --- | --- |
|  | unadjusted OR<br>(95% CI) | *p* value | adjusted OR<br>(95% CI) | *p* value | unadjusted OR<br>(95% CI) | *p* value | adjusted OR<br>(95% CI) | *p* value | unadjusted OR<br>(95% CI) | *p* value | adjusted OR<br>(95% CI) | *p* value |
| A type | 0.83(0.30, 2.32) | 0.727 | 1.31(0.38, 4.53) | 0.671 | 0.74(0.22, 2.52) | 0.632 | 1.19(0.29, 4.90) | 0.809 | 0.88(0.27, 2.89) | 0.826 | 0.79(0.19, 3.20) | 0.741 |
| B type | 1.38(0.44, 4.29) | 0.583 | 2.12(0.55, 8.15) | 0.275 | 1.07(0.31, 3.68) | 0.920 | 1.89(0.43, 8.18) | 0.398 | 1.07(0.31, 3.68) | 0.920 | 1.20(0.28, 5.13) | 0.814 |
| O type | 1.00 | - | 1.00 | - | 1.00 | - | 1.00 | - | 1.00 | - | 1.00 | - |

ARDS, Acute respiratory distress syndrome; AKI, Acute kidney injury; CI, confidence interval; OR, odds ratio; ORs and *p* values were determined for the comparison of blood type A or B with blood type O through logistic regression using backward elimination. Adjusted ORs for age, sex, and medical history.

## Discussion

The current retrospective investigation found that blood type A individuals were more sensitive to SARS-CoV-2. Although patient age was a factor of mortality in the present study as reported in previous studies [1], blood type distribution was not a relevant factor of ARDS, AKI, and mortality in COVID-19 patients. To our knowledge, this is the first study to report a conceivable association between ABO blood types and ARDS, AKI, and death risk, and to demonstrate that ABO glycobiology may not play a crucial role in the development of COVID-19. We believe that these findings are indicative since it probed the relationship between blood types and ARDS, AKI, and mortality, in addition to susceptibility or age-related mortality in COVID-19 sufferers.

COVID-19 now posts a great threat to human health worldwide. Thus, global efforts are being made to fight against this serious virus infection. Recently, scholars have observed that age and gender are correlated with SARS-CoV-2 infection susceptibility or severity [1]. What we found in this study provided some new information from the blood type distribution angle in the mild and critical cohorts. ARDS, AKI, and mortality were not correlated with blood type distribution in COVID-19 patients. Our observations partially clarified the issues regarding the blood type discrimination in the COVID-19 fighting battle. Since blood type is not risk factor for ARDS, AKI, and mortality, average people and health care workers should not overestimate the genetic susceptibility by placing a certain blood group at risk for poor prognosis. This kind of research methods also may be useful for evaluating the relation between blood type and other diseases, such as West Nile virus infection, hepatitis C, diabetes mellitus, and so on. The susceptibility of different ABO blood groups to these diseases has also been investigated [24, 27, 28], while the progression or prognosis has not been analyzed yet.

The correlation between age, gender, and the susceptibility of SARS-CoV-2 has been investigated in the past several months. What we observed in the current study also confirmed that the aging population are vulnerable to SARS-CoV-2 infection. Consequently, specific preventive measures and therapies should be implemented in the aging population. In addition, the blood type A population was also more susceptible to SARS-CoV-2 infection, which was consistent with the results of previous similar investigations on West Nile virus infection and plasmodium falciparum susceptibility [27, 29]. Although the sample in our study was relatively small, the data used in the study were obtained from multiple medical centers and may better represent this patient group. Further large-scale investigations are needed to resolve this concern.

Previous reports found that the blood group O population was less susceptible to SARS virus [2], while we did not obtain the same conclusion in the study on novel coronavirus. Instead, our study demonstrated that blood group A individuals were more susceptible to SARS-CoV-2 infection. We speculate that the main reason is that specific viral protein structure will lead to differences in susceptibility, since the unique virus gene and encoded structure have been resolved [30]. Furthermore, the previous study was aimed at investigating an individual’s inclination to SARS virus infection and was conducted on 45 younger healthcare workers who were exposed to an index SARS patient [2]. It is difficult to determine why the blood type A population is more susceptible to SARS-CoV-2 virus. Actually, in most situations, why some diseases occur in a specific blood type population is not yet clear, except that Duffy blood type defects determine the malaria infection in Africa. Apart from the regular expression on the red blood cell surface, blood group antigens are also widely distributed on various cells and sometimes in body fluids, including respiratory epithelial cells and alveolar epithelial cells. We believe that the genetic susceptibility of blood type glycophorin may function via receptor-mediated affinity binding, especially in the invasion mechanism. Blood group antigens are proven to be effective receptors for several infectious microorganisms [31]. The specific ABO glycan antigen receptors binding to the virus spike (S) protein may support virus entry during infection.

Given the effects on inflammation, endothelial function, and microvascular coagulation of ABO glycoproteins by changing blood concentrations of soluble ICAM-1, selectins, vWF, and thrombomodulin [32-34], the ABO glycans may be important mediators of ARDS or AKI in critically ill patients. Previous studies showed that blood type A was associated with a higher risk of developing ARDS and AKI in the ICU among individuals of European descent, but not in those with African descent [34, 35]. This means that racial divergence and environmental factors may alter the ABO-ARDS or ABO-AKI associations. It is plausible to hypothesize that the existing conclusion drawn in European descent with trauma or severe sepsis may not be applied to the Asian population with COVID-19. Furthermore, there is insufficient evidence regarding the pathophysiologic mechanism and features of ARDS or AKI that occurs secondary to SARS-CoV-2 infection [32-35].

The present study has several limitations. First, it is not feasible to use a prospective cohort to determine when patients with mild symptoms progress to the critically ill stage, since the epidemic situation was very serious at that time and all the resources were in used for emergency care. Second, there are no existing wild cohorts with available demographic and clinical characteristics that can be used as a reference group because patients who developed fever without SARS-CoV-2 infection were not identified during the corresponding period. Thus, it is impossible to analyze the correlation between blood type distribution and COVID-19 using a multiple-factor model logistic model. We had to use the chi-square test and calculate the OR value to provide an estimate. This strategy cannot rule out the possibility that blood type is the only factor associated with SARS-CoV-2 infection and progression. Our observations were only based on the results of the epidemic data analysis. The biological evidence between blood type and SARS-CoV-2 susceptibility and progression requires further cellular and molecular investigation.

The current retrospective study of two cohorts confirmed that blood type A populations are more susceptible to SARS-CoV-2 infection. However, the outcomes including ARDS, AKI, and death are poor irrespective of ABO blood group distribution in critically ill patients with COVID-19, although blood type O sufferers are unlikely to progress into the critical stage after SARS-CoV-2 infection. Additionally, our study identified that age is a risk factor for the development of ARDS or death in critically ill patients with COVID-19. These findings are significant since it deeply probed the correlation between blood type and progression or prognosis in addition to susceptibility. With this kind of research, we will be able to support the fight against COVID-19 around the world.

## Data Availability

All data generated or analyzed during this study are included in this article.

